# Bradykinin Measurement by LC-MS/MS in Hereditary Angioedema Subjects Enhanced by Cold Activation

**DOI:** 10.1101/2024.12.23.24319547

**Authors:** Jinguo Chen, Yunkou Wu, Joseph Chiao, J. Joanna Yu, Jing Yu, Mark D. Scarupa, Lili Wan, H. Henry Li

## Abstract

**Background:** Bradykinin (BK) is a key mediator responsible for swelling episodes in hereditary angioedema due to C1INH deficiency/dysfunction (HAE-C1INH). Current BK measurement faces many challenges primarily related to very low levels and instability, and is not feasible for application in clinical settings.

**Objective:** This study aimed to develop a novel method to overcome the issues in current protease inhibitor-based methods for measuring endogenous BK and its metabolites.

**Methods:** The blood from HAE-C1INH and healthy volunteers were collected and subjected to cold activation for contact system. Cold-induced BK and its major metabolites were measured via Liquid Chromatography-Tandem Mass Spectrometry (LC-MS/MS). The protocol was established according to the US FDA bioanalytical validation guidelines as a CLIA laboratory-developed test. The BK measurement was optimized based on blood sample types, collection methods, time windows, and temperature/storage conditions.

**Results:** EDTA whole blood samples without protease inhibitors incubated at 4 degree for 1 to 3 days produced over 100-fold differences in total BK levels between HAE-C1INH subjects and healthy volunteers (324.3 +/- 54.7 ng/mL, n=33; vs 2.3 +/- 0.3 ng/mL, n = 43; mean +/- S.E.M., p < 0.001). The sensitivity and specificity were 90.9% and 97.1% respectively. BK levels highly correlated with the plasma kallikrein activity in the same samples.

**Conclusions:** Whole blood under cold activation demonstrated remarkable elevation of BK levels in HAE-C1INH subjects, while minimally affecting healthy individuals. The assay has validated accuracy, precision and stability. It may serve as a reliable and robust tool for HAE diagnosis and management.

**Clinical Implication:** Cold-induced Bradykinin measurement can be used as a new biomarker for diagnosis, disease monitoring and guiding therapeutic options for HAE-C1INH and other bradykinin-mediated angioedema (AE-BK), with or without identifiable genetic mutations.

## Introduction

Bradykinin (BK) is a mediator involved in multiple pathophysiological processes.^1^ It is primarily produced through the cleavage of high-molecular-weight kininogen by activated plasma kallikrein (PK). Two receptors (B1R and B2R) are specific for BK.^2^ BK binding to B2R on vascular endothelium may lead to vasodilation and increased vascular permeability. BK is the key mediator for angioedema attacks in hereditary angioedema due to C1INH deficiency (HAE-C1INH).^3,4^ BK is rapidly metabolized in plasma by a few peptidases. The intact form of BK is BK1-9 (RPPGFSPFR). Its major metabolites include BK1-8, BK1-7, BK1-5, and BK2-9.^5^ Recent studies indicated that almost no breakdown product of BK is functionally inactive.^2^

Immunoassays for BK measurements date back to 1979.^6–8^ It’s less commonly used today due to significant cross-reaction with similar peptides. Liquid chromatography coupled with tandem mass spectrometry (LC-MS/MS) can overcome immunoassay-related limitations. LC-MS/MS also has the advantage of shorter development times, greater accuracy and precision. It can also multiplex and readily distinguish between closely related analogs, metabolites, or endogenous interferences.^9^ Accurate quantification of BK can still be challenging due to its low baseline levels (low pg/mL), short plasma half-life (<30 seconds), and variability related to blood collection, processing, and proteolytic activities.^2^

Using protease inhibitor (PI) cocktails to stabilize BK after blood collection could improve the analytical sensitivity and repeatability.^10^ Several LC-MS/MS methods have been successfully developed to detect endogenous BK levels.^9,11^ However, the reported BK levels varied from 0.2 ng/mL to 160 ng/mL in healthy volunteers.^11,12^ In one study, although the sum of BK levels in HAE-C1INH (18.0 ± 6.3 pmol/L, mean ± S.E.M.) was higher than healthy volunteers, 7 out of 9 HAE-C1INH patients had BK values close to upper range of normal, indicating a very narrow diagnostic window.^13^ The broad range of reported normal levels, and very narrow separation between HAE-C1INH and normal subjects would limit its clinical application.

The blood contact system is more unstable and easily activated at 4°C *ex vivo* in HAE-C1INH subjects but not in normal controls.^14,15^ We deduced that BK levels could have similar changes. In this study, we modified previously reported LC-MS/MS methods for BK measurement^11,16^ and applied cold activation during blood sample preparation. EDTA whole blood was collected without PI and the contact system was fully activated in the cold for 1-3 days before BK analysis with LC-MS/MS methods. The results showed that the cold-induced total BK peptides increased more than 100-fold in HAE-C1INH patients compared to healthy volunteers, indicating a robust diagnostic value in HAE disease.

## MATERIALS AND METHODS

### Human subjects

Blood samples were collected from HAE-C1INH patients confirmed by previous C4, C1INH, and SERPING1 genetic tests. HAE-C1INH subjects on long-term prophylaxis were excluded. Healthy volunteers (HV) were also recruited to establish the reference interval and for method validation. HV excluded subjects with any forms of angioedema and recent respiratory illness (COVID-19, etc.). Concurrent use of angiotensin-converting enzyme inhibitors (ACEIs) or estrogen-containing birth control pills were also excluded. Informed consent was obtained from each subject recruited through the Angioedema Centers of Reference and Excellence at the Institute for Asthma and Allergy (MD, USA). The study was conducted following the guidelines of a clinical study protocol approved by a central Virant Institutional Review Board (IRB protocol number: Virant-A0001).

### Blood sample collection and processing

EDTA or citrate whole blood or plasma samples were collected for analysis. The samples were aliquoted 0.1mL per aliquot into Eppendorf tubes and incubated at room temperature or 4°C at intervals ranging from 0 hours to 5 days. At the specified time points, these samples were mixed with 0.5 ml of 80% ethanol, vigorously vortexed, and kept at room temperature, 4°C, -20°C, or -80°C until analysis. The ethanol-mixed samples were then briefly centrifuged, and the resulting supernatants were carefully transferred to new Eppendorf tubes for the sample extraction step.

### Sample Extraction with C18 SPE Cartridge

A stable isotope-labeled internal standard (IS) mix containing BK1-5 (¹³C/¹⁵N) (Innovagen, Lund, Sweden) and [Phe8Ψ(CH-NH)-Arg9]-BK (Tocris Bioscience, Minneapolis, MN, USA) was spiked into each ethanol-mixed sample. These samples were dried using a SpeedVac Vacuum Concentrator at 70°C until solid pellets were obtained. The dried samples were then reconstituted with 0.1% formic acid, which was subsequently processed through solid-phase extraction (SPE).

SPE was carried out on ResPrep C18 extraction cartridges (Restek, Bellefonte, PA, USA), featuring a 1 ml volume and 100 mg beds. All samples were loaded onto a preconditioned cartridge, then washed with 0.1% formic acid, and followed by 80% ethanol elution. The ethanol in the eluates was evaporated using SpeedVac at 70°C until dry. The dried samples were dissolved in 100 μL of a solution containing 0.3% trifluoroacetic acid in a mixture of acetonitrile and water (25/75, v/v) for subsequent LC-MS/MS analysis.

### LC-MS/MS Method Development

LC-MS/MS analysis was performed using a SCIEX (Framingham, MA, USA) Triple Quad 5500+ QTRAP MS/MS system coupled with a Shimadzu Nexera 40 HPLC system. A Kinetex Biphenyl column with a guard column (Phenomenex, Torrance, CA) was employed for chromatographic separation. Optimal separation was achieved using a gradient elution program, with the specific conditions detailed in **Table E1** in the Online Repository available at www.jaci-global.org.

The mass source of the spectrometer was carefully fine-tuned for optimal ionization and efficient ion transfer. The spray voltage was set at 1500 V, curtain gas at 35 PSI, collision gas at 8 PSI, Ion Source Gas 1 at 40 PSI, and Gas 2 at 45 PSI, respectively. The vaporizer temperature was consistently maintained at 300°C. Peptide-specific parameters and the instrument settings are detailed in **Table I**.

MS data were acquired using Analyst software v1.7.3 (AB Sciex), and analyte results were quantified and reported with Sciex OS v3.0.0.3339 (AB Sciex). Eight levels of working calibrators, with concentrations ranging from 0.1 to 1000 ng/mL, as well as two levels of quality control (QC) samples (QC low and QC high), were used in this study. Additionally, an IS mixture including ¹³C/¹⁵N-labeled BK1-5 (for BK1-5) and Phe8Ψ(CH-NH)Arg9 BK (for BK1-9, BK1-8, and BK1-7), were added during calibrator and sample preparation. Sample concentrations were normalized with the IS and calculated from the standard curves.

Although detectable, BK2-9 has been shown to be consistently low or below detectable limit and thus was subsequently excluded from our analysis. In this study total BK only includes BK1-9, BK1-8, BK1-7 and BK1-5.

### LC-MS/MS Method Validation

Following the US FDA bioanalytical validation guidelines^17^ and in compliance with CLIA regulations for a laboratory-developed test (LDT), we conducted linearity, accuracy, precision, sensitivity, recovery, matrix effect, carryover, and stability assessment. The specific validation designs were described in the Online Repository available at www.jaci-global.org. The results of these validation experiments met the assay acceptance criteria before being accredited by the College of American Pathologists (CAP) and being added to the clinical test menu.

### Plasma Kallikrein amidolytic activity assay for method comparison

The PK amidolytic activity assay is an colorimetric method that utilizes the chromogenic substrate H-D-Pro-Phe-Arg-p-nitroanilide (Bachem, Torrance, CA, USA), following protocols previously described with modifications.^18^ To ensure the specificity of the assay for PK activity and to eliminate interference from Factor XIIa (FXIIa), a specific inhibitor, PKSI-527, was employed to inhibit FXIIa activity. For the method comparison procedure, the PKa assay was conducted alongside the BK LC-MS/MS assay, with both sample sets undergoing cold incubation at similar time points before analysis. This parallel evaluation was designed to validate the consistency and reliability of the BK LC-MS/MS method in detecting contact system activation and to help establish its diagnostic utility for bradykinin-related disorders.

## RESULTS

### LC-MS/MS Method Development

#### HPLC separation column

A Kinetex Biphenyl column (50 mm × 1 mm, 5 μm) was selected and utilized in this study. The optimized separation conditions were shown in **Table E1** in the Online Repository. Kinetex Biphenyl is a high-efficiency core-shell product capable of adding extra separation power to the analysis of non-polar and polar compounds.^19^

#### Mass spectrometry conditions

During method development, different multiple-charged precursors and fragments were monitored. The two most intense and selective ones for each analyte were chosen in this study (**Table 1**). Fragment 1 was for quantitative analysis and fragment 2 was for confirmatory purposes. Many peptides produce intense fragments below *m/z* 200 and result in a high background in extracted samples.^9^ In this method, the use of highly specific ion fragments with high m/z values (>400 for Fragment 1) and scheduled “Multiple Reaction Monitoring Pro Algorithm” for MS signal acquisition yielded significantly improved specificity and accuracy (**Figure E1** in the Online Repository).

**Table 1.** MS/MS settings for BK Peptides and Internal Standards (IS)

| Transition (m/z) | Retention Time (min) | IonDrive | DP* (volts) | EP* (volts) | CE* (volts) | CXP* (volts) |
| --- | --- | --- | --- | --- | --- | --- |
| 530.9→522.2 | 3.93 | Bradykinin1-9 1 | 40 | 13 | 33 | 32 |
| 530.9→904.3 | 3.93 | Bradykinin1-9 2 | 100 | 8 | 33 | 32 |
| 452.8→642.4 | 4.39 | Bradykinin1-8 1 | 75 | 5 | 22 | 40 |
| 452.8→263.2 | 4.39 | Bradykinin1-8 2 | 85 | 13 | 22 | 16 |
| 379.2→642.6 | 3.59 | Bradykinin 1-7 1 | 60 | 15 | 18 | 40 |
| 379.2→555.5 | 3.59 | Bradykinin 1-7 2 | 70 | 7 | 18 | 44 |
| 523.8→515.1 | 3.74 | Phe8Ψ(CH-NH)Arg9 BK IS | 120 | 8 | 33 | 20 |
| 287.2→408.2 | 3.15 | Bradykinin1-5 1 | 70 | 12 | 16 | 26 |
| 287.2→320.2 | 3.15 | Bradykinin1-5 2 | 65 | 14 | 14 | 22 |
| 292.2→408.2 | 3.15 | Bradykinin1-5 IS | 55 | 14 | 18 | 26 |
| 452.8→404.4 | 4.25 | Bradykinin2-9 1 | 100 | 9 | 28 | 20 |
| 452.8→807.4 | 4.25 | Bradykinin2-9 2 | 100 | 7 | 28 | 24 |

\* DP (declustering potential), EP (entrance potential), CE (collision energy), and CXP (collision cell exit potential)

#### Blood sample preparation and cold activation

Our initial studies revealed that using protease inhibitors to capture BK peptides produced *in vivo* for MS measurement was not a reliable approach, largely due to the narrow quantitative thresholds observed between samples from healthy individuals and those with HAE-C1INH. Previous research demonstrated that *ex vivo* cold incubation of blood samples activates the contact system in most HAE patients, evidenced by increased plasma kallikrein (PK) and Factor XII (FXII) enzymatic activity.^15,20^ We optimized the cold incubation protocol and examined BK levels in six healthy control and five HAE-C1INH patients, comparing their samples under cold-incubated vs. room-temperature conditions for 24 hours. As shown in **Figure 1**, cold-incubation drastically enhanced the BK levels in five HAE-C1INH patients, while minimally affecting six healthy controls, indicating that an enhanced (or widened) separation window of these two groups can be achieved under cold-activation conditions.

**Figure 1.**
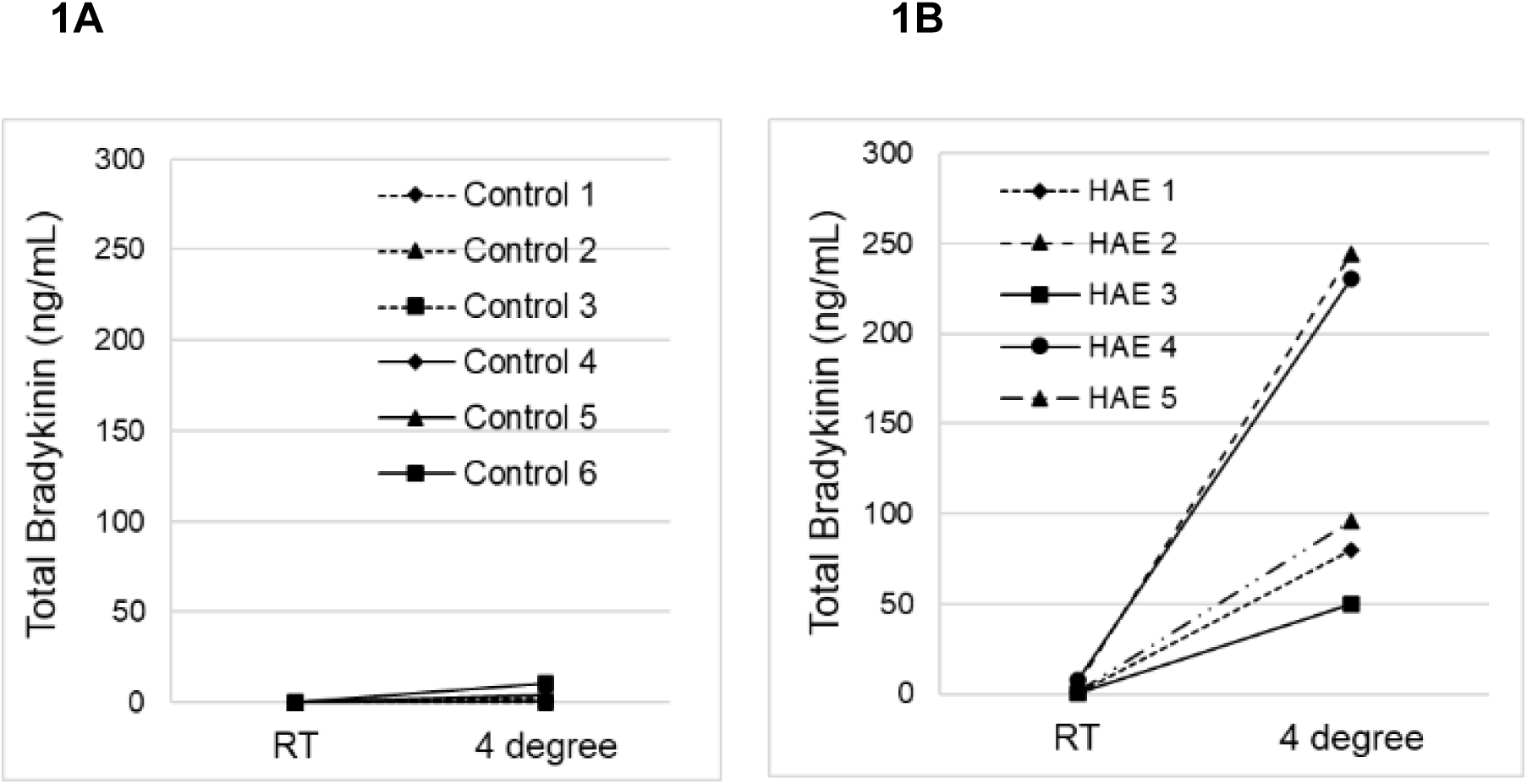
Cold activation preferably produced high levels of BK in HAE-C1INH patients. The validated LC-MS/MS method was utilized for BK measurements of EDTA whole blood samples from six healthy controls (HV) and five HAE-C1INH patients incubated at room temperature (RT) or at 4 °C for 24 hours. (**1A**) No significant differences were observed in HV between RT and 4°C. (**1B**) In HAE-C1INH subjects, the BK levels showed great differences between RT and 4°C incubation.

To minimize the manipulation of blood samples and to avoid inducing more spontaneous activation, whole blood was identified as the preferred sample type in this study. Both plasma and serum artificially increased the levels of BK and its metabolites (Total BK) as compared with whole blood after cold incubation (**Figure 2A**). Among anticoagulants used in blood collection devices, EDTA was favored over citrate, as our data demonstrated that 24-hour cold incubation consistently yielded high BK and metabolite levels in EDTA-treated blood but not in citrate-treated samples (**Figure 2A).** We also examined the difference of BK measurements between the first and second peripheral blood collection tubes from five subjects because the literature suggests that BK levels may be affected by torniquet use and the pain related to the blood collection. However, based on our analysis, no significant differences in quantitative BK values were observed between first and second sample sets when cold incubation were applied (**Figure 2B**).

**Figure 2.**
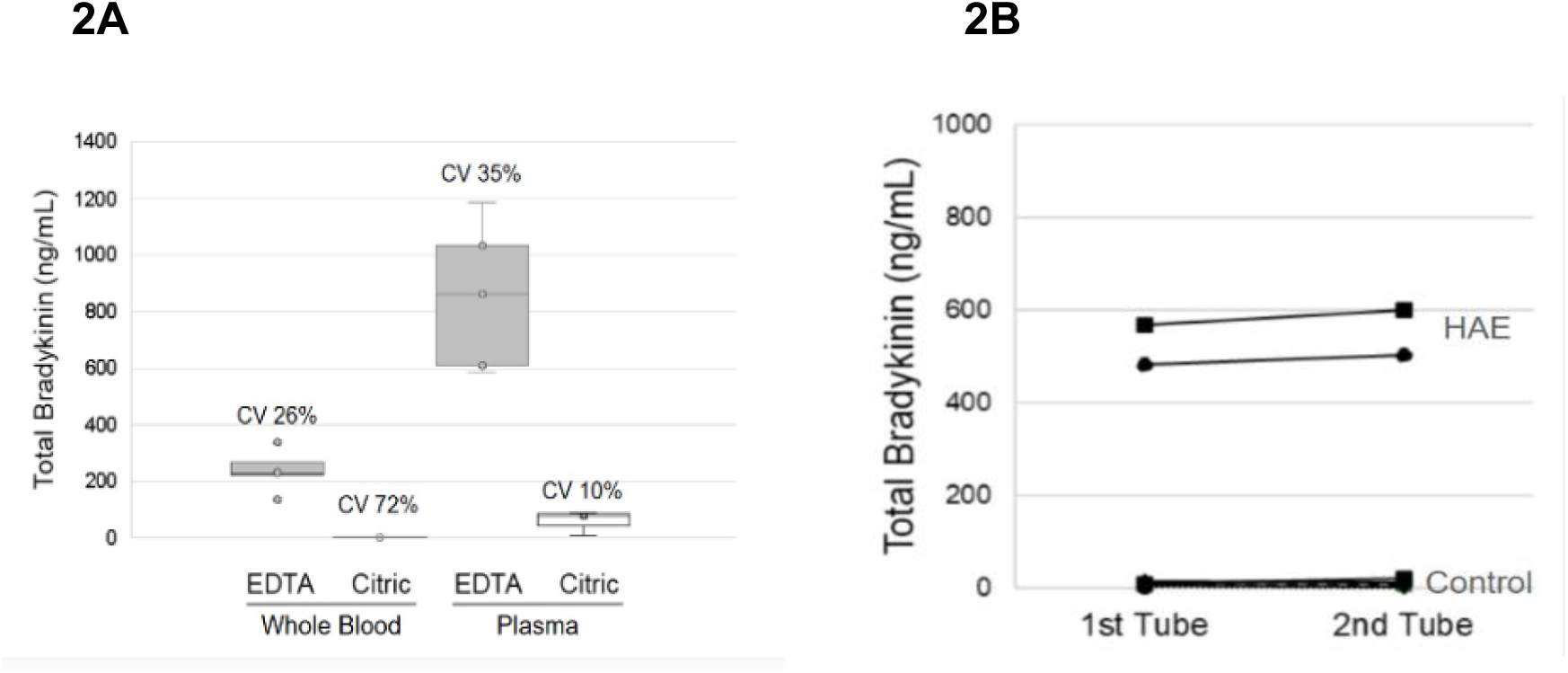
Optimization of specimen selection. (**2A**): Plasma artificially increased the levels of BK and its metabolites (Total BK) after cold incubation for 24 hours as compared with whole blood in the 10 paired samples from five subjects. Cold incubation consistently yielded total BK levels in EDTA-treated blood but not in citrate-treated samples. (**2B**): There were no significant differences of total BK levels in the first and the second EDTA blood collection tubes.

To assess the temporal dynamics of BK and its metabolites after cold activation, we monitored samples over a 5-day period. Total BK levels in a representative HAE-C1INH subject increased from 13.9 ng/mL on day 0 to a peak of 276.0 ng/mL on day 1, subsequently stabilizing between 204.4 and 226.2 ng/mL on days 3 and 5 respectively. During this period, BK1-9 and BK1-8 levels declined, while BK1-7 and BK1-5 levels continued to rise. However, the distribution profiles of individual BK metabolites varied among subjects (**Figure 3A**). Most subjects exhibited peak total BK levels between day 1 and 3, prompting us to examine BK levels at both time points in this study, with the higher value from the two time points reported.

**Figure 3.**
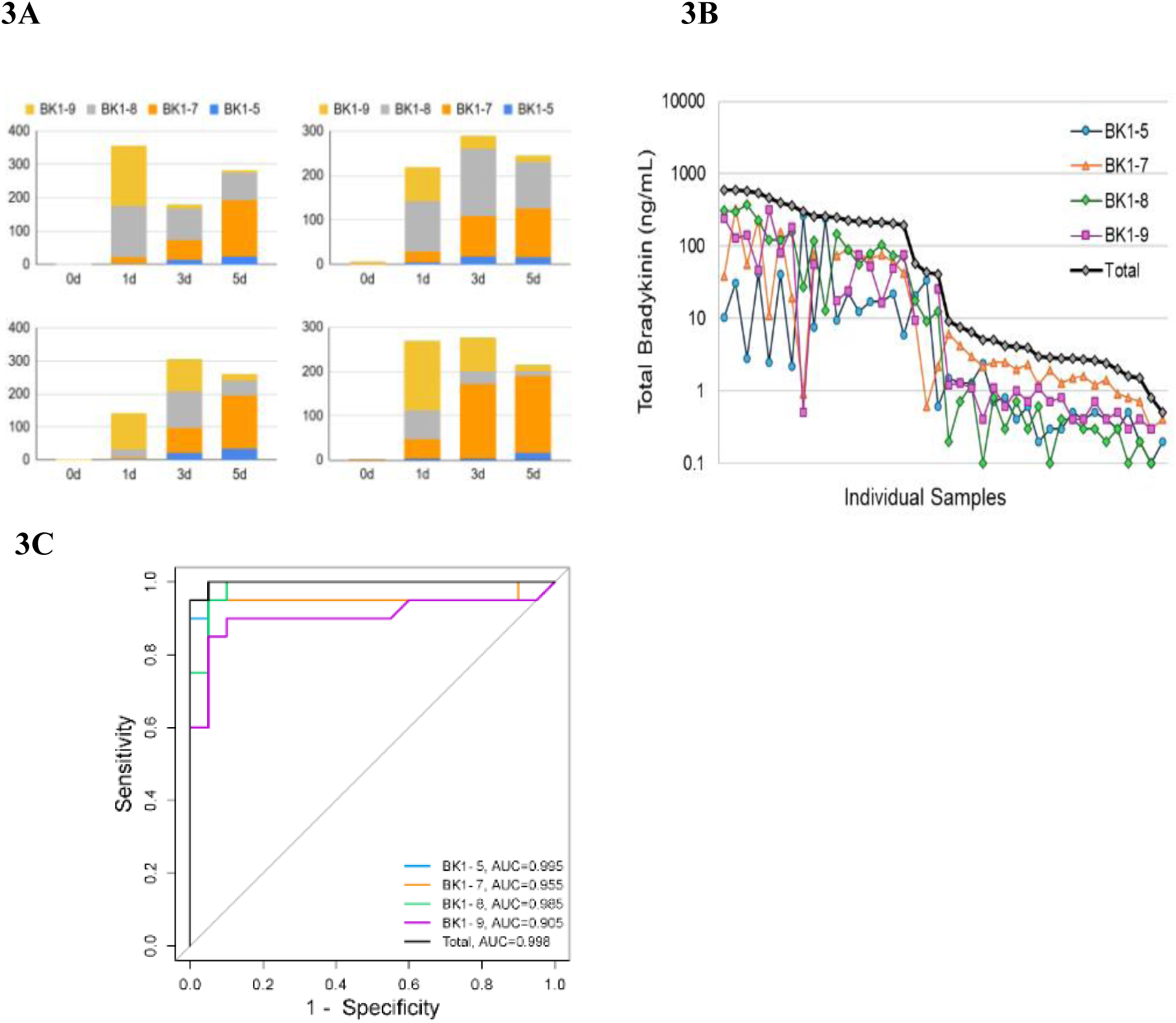
Total BK delivers better diagnostic performance in HAE-C1INH than individual peptides. (**3A**) The distribution profiles of individual BK metabolites varied among subjects. Most subjects exhibited peak total BK levels between day 1 and 3. (**3B**): No repeated pattern of individual BK metabolite was found in HAE-C1INH samples. The plot of the total BK values was a better reflection of the cold-activated samples. (**3C**) Receiver operating characteristic (ROC) curve analysis (20 HAE-C1INH and 20 HV) showed that total BK had the highest area under the curve (AUC), indicating the best diagnostic value for HAE-C1INH.

Ethanol was used to stop cold activation by denaturing enzymatic proteins. To evaluate the stability of ethanol-treated samples, we stored cold-activated, ethanol-precipitated EDTA whole blood at 4 °C or -80 °C for varying durations before analysis. BK and its metabolites remained stable for up to four weeks at 4 °C and up to eight months at -80 °C. A Levey-Jennings chart (**Figure 4**) showed that total BK concentrations varied within ±15% of expected values, confirming the stability of ethanol-treated samples.

**Figure 4.**
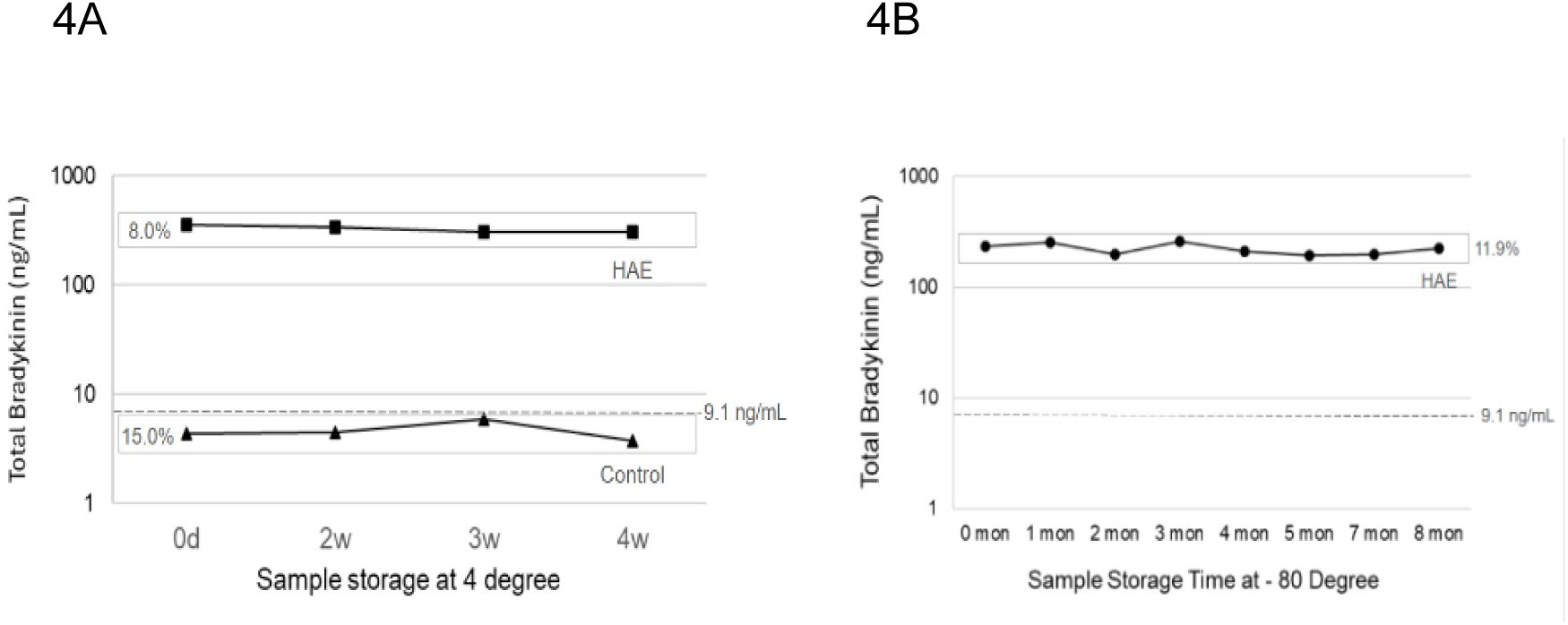
Great blood sample stability after cold activation. Ethanol was used to stop cold activation by denaturing enzymatic proteins. BK and its metabolites in ethanol-treated blood remained stable for up to four weeks at 4 °C (**4A**) and up to eight months at -80 °C (**4B**).

Our specimen collection and processing experiments validated that EDTA whole blood should be collected, aliquoted, and subjected to cold incubation for 1 to 3 days post-collection. Ethanol was then applied to stop cold activation before the chromatography step, which ensured reliable and reproducible BK measurements.

### LC-MS/MS Method Validation

#### Linearity

Five replicates of eight calibrators were prepared to cover the desired concentration range of 0.1 ng/mL to 1000 ng/mL. The calibrators were then analyzed using the instrument method described above. The concentrations and measurement accuracy of the analytes were calculated using the Sciex OS software through linear regression. The linearity of calibration curves for all analytes displayed coefficients of correlation values surpassing 0.985 (**Table E2** in the Online Repository). In addition, all analytes exhibited exceptional accuracy with relative errors below 15%.

#### Accuracy and Precision

Four levels of quality control samples were prepared to assess within-run accuracy and precision, as well as between-run precision. These samples included an LLOQ (0.1 ng/mL), QC1 (1 ng/mL), QC_med_ (10 ng/mL) and QC2 (70 ng/mL). The samples were injected in five batches over three days to evaluate precision and accuracy. These assessments consistently demonstrated that both within-run and between-run precision were maintained below 15% relative errors, while accuracy remained within the acceptable range of ± 15% of the nominal concentrations (**Table 2**) as stipulated in the FDA validation guidelines.^17^

**Table 2.** Accuracy and precision validation results.

| Analytes | QC Sample | Nominal value (ng/ml) | n | Accuracy (RE%) |  |  |  | Precision (CV%) |  |
| --- | --- | --- | --- | --- | --- | --- | --- | --- | --- |
|  |  |  |  | Day1 | Day2 | Day3 | Inter-run | Intra-run | Inter-run |
| BK1-5 | LLOQ | 0.1 | 15 | -0.2 | 10.0 | 11.0 | 6.9 | 3.1 | 5.1 |
|  | QC1 | 1.0 | 15 | 2.6 | 9.1 | -0.9 | 3.6 | 3.5 | 4.0 |
|  | QCmed | 10.0 | 15 | 0.5 | 0.1 | -3.9 | -1.1 | 0.8 | 0.4 |
|  | QC2 | 70.0 | 15 | -1.0 | 0.1 | 0.8 | -0.1 | 0.3 | 0.7 |
| BK1-7 | LLOQ | 0.1 | 15 | 8.0 | -1.0 | 11.0 | 6.0 | 2.9 | 4.8 |
|  | QC1 | 1.0 | 15 | 2.6 | 9.1 | -0.9 | 3.6 | 3.5 | 4.0 |
|  | QCmed | 10.0 | 15 | 5.0 | 7.5 | 0.3 | 4.3 | 2.3 | 2.9 |
|  | QC2 | 70.0 | 15 | 0.7 | 5.5 | 0.9 | 2.4 | 1.9 | 2.6 |
| BK1-8 | LLOQ | 0.1 | 15 | 9.0 | -0.5 | 15.0 | 7.8 | 3.9 | 7.9 |
|  | QC1 | 1.0 | 15 | 0.8 | 8.8 | 1.1 | 3.6 | 3.3 | 3.6 |
|  | QCmed | 10.0 | 15 | 9.5 | 2.9 | -5.4 | 2.3 | 2.5 | 6.0 |
|  | QC2 | 70.0 | 15 | 4.1 | 5.8 | -6.4 | 1.2 | 2.9 | 5.0 |
| BK1-9 | LLOQ | 0.1 | 15 | 2.0 | -12.0 | -6.0 | -5.3 | 6.1 | 10.7 |
|  | QC1 | 1.0 | 15 | -0.9 | 3.5 | -0.2 | 0.8 | 2.2 | 6.8 |
|  | QCmed | 10.0 | 15 | -3.5 | 2.6 | -2.5 | -1.1 | 2.1 | 3.3 |
|  | QC2 | 70.0 | 15 | -0.7 | 4.0 | -0.5 | 0.9 | 2.0 | 2.1 |

QC: quality control; LLOQ: lower limit of quantification; RE: relative error; CV: coefficient of variation

#### Sensitivity and Specificity

The Lower Limit of Quantification (LLOQ) determined the sensitivity of the method which was set at 0.1 ng/mL for all kinin peptides. The LLOQ is often evaluated by signal-to-noise ratio (S/N), with an acceptable S/N ratio being larger than 10. All analytes at LLOQ consistently displayed S/N values exceeding 10 (**Table E2** in the Online Repository), meaning that the LLOQ could be set lower than 0.1 ng/mL. Although the analytical sensitivity of our method was slightly lower than previously reported in the literature^11,12^, it exceeds the threshold to detect all kinin peptides at ng/mL level after cold activation.

To evaluate assay specificity, common over-the-counter medications and chemicals including cotinine, nicotine, acetaminophen, caffeine, ibuprofen, Naproxen at 25 mg/mL and phentermine, pseudoephedrine at 2.5 mg/mL were added to blank matrix samples, along with known concentrations of the analytes. The concentrations of each analyte were then measured in the presence and absence of the interfering substances. The results showed that the interference effect from the added substances was less than 10% for all analytes, indicating minimal impact on the high specificity of this method.

#### Carryover

Carryovers were evaluated by measuring solvent blank injected after injection of the highest calibrator (Calibrator Level 8, 1000 ng/mL). The acceptable carryover of analytes will have a peak area of less than 20% of the lowest calibrator (Cal1, 0.1 ng/mL). The results demonstrated that the Area_Blank_/Area_Cal1_(%) was less than 20% for all analytes and two internal standards, therefore no carryover was observed.

#### Matrix Effect

Matrix effect (*ME*_ionization_) refers to interferences that can cause ion suppression or ion enhancement in the mass signal due to the matrix composition. Post-extraction matrices from five healthy donors were spiked with calibrators at low and high QC levels to assess the matrix effect. The measured concentrations obtained from these matrices were then compared to the ones obtained from the neat solvent. The results demonstrated that all analytes fell within the acceptable range of 50% to 100%, and within the acceptable range of 50% to 150%, indicating that the matrix composition did not significantly affect ionization.

#### Calibrator stability

Calibrator sample stability was assessed across three concentration levels: QC1 (1 ng/mL, low), QCmed (10 ng/mL, medium), and QC2 (70 ng/mL, high). The calibrator samples were stored under room temperature, refrigeration (2-8 °C), and freezer (-20 °C) conditions. The samples were extracted and measured on days 1, 3, 5, 7, 10, and 14, with the acquired results compared against those from day 0. The results showed less than 15% changes for all analytes after storage at different conditions, which meets the acceptance criteria.

Validation studies confirmed that BK measurement with LC-MS/MS is a reliable, acceptable method with an accuracy of 97%, precision of >99%, analytical sensitivity of 0.1 ng/mL, analytical specificity of >99% and reportable range of 0.1 ng/mL to 1,000 ng/mL (for each BK metabolite).

### Diagnostic value of cold-induced BK assay in HAE-C1INH

#### Total BK *vs.* individual peptides

BK1-9, BK1-8, BK1-7, and BK1-5 were simultaneously measured in this study. Each patient displayed different patterns for individual BK peptides (**Figure 3A**) after cold activation. BK1-9 seemed to degrade into 1-8, 1-7 and 1-5 with the degradation rate varying from subject to subject. The BK metabolites and total BK values from 33 patients with HAE-C1INH found no repeated pattern of BK metabolite value (**Figure 3B**). However, the plot of the total BK values was a better reflection of the cold-activated samples. Receiver operating characteristic (ROC) curve analysis (20 HAE-C1INH and 20 HVs) in **Figure 3C** showed that total BK has a highest AUC (area under the curve), indicating the best diagnostic value for HAE-C1INH. We thus chose the total BK (the sum of BK1-9, BK1-8, BK1-7 and BK1-5) for final reporting. Reference interval of total BK from 43 healthy volunteers was determined as 0.1 to 9.1 ng/mL.

#### Comparison of BK levels with PK activity

It has been demonstrated that plasma kallikrein (PK) initiates and plays the major role in cold activation of the contact system.^15^ Activated PK cleaves the HMWK and produces the BK. Therefore, PK amidolytic activity (PKa) was used for method comparison in this study. In a validation cohort, both BK levels and PK activities were significantly increased after cold activation in 33 HAE-C1INH patients **(Figures 5A)** compared with those in 43 healthy volunteers. The differences in total BK levels are more than 100-fold on average (HAE 324.3 ± 54.7 ng/mL vs healthy volunteers 2.3 ± 0.3 ng/mL; mean±S.E.M., p < 0.001). The PK activities showed a strong correlation with the BK levels in the same samples. Nearly all samples demonstrated changes in the same direction across control groups **(Figures 5B)** and HAE (**Figures 5C)**. Receiver Operating Characteristic (ROC) analysis highlighted strong diagnostic performance for both BK and PKa assays **(Figure 5D)**. With the normal cutoff for the BK assay set at 9.1 ng/mL, the sensitivity was 90.9%, and the specificity was 97.1% for HAE-C1INH diagnosis.

**Figure 5.**
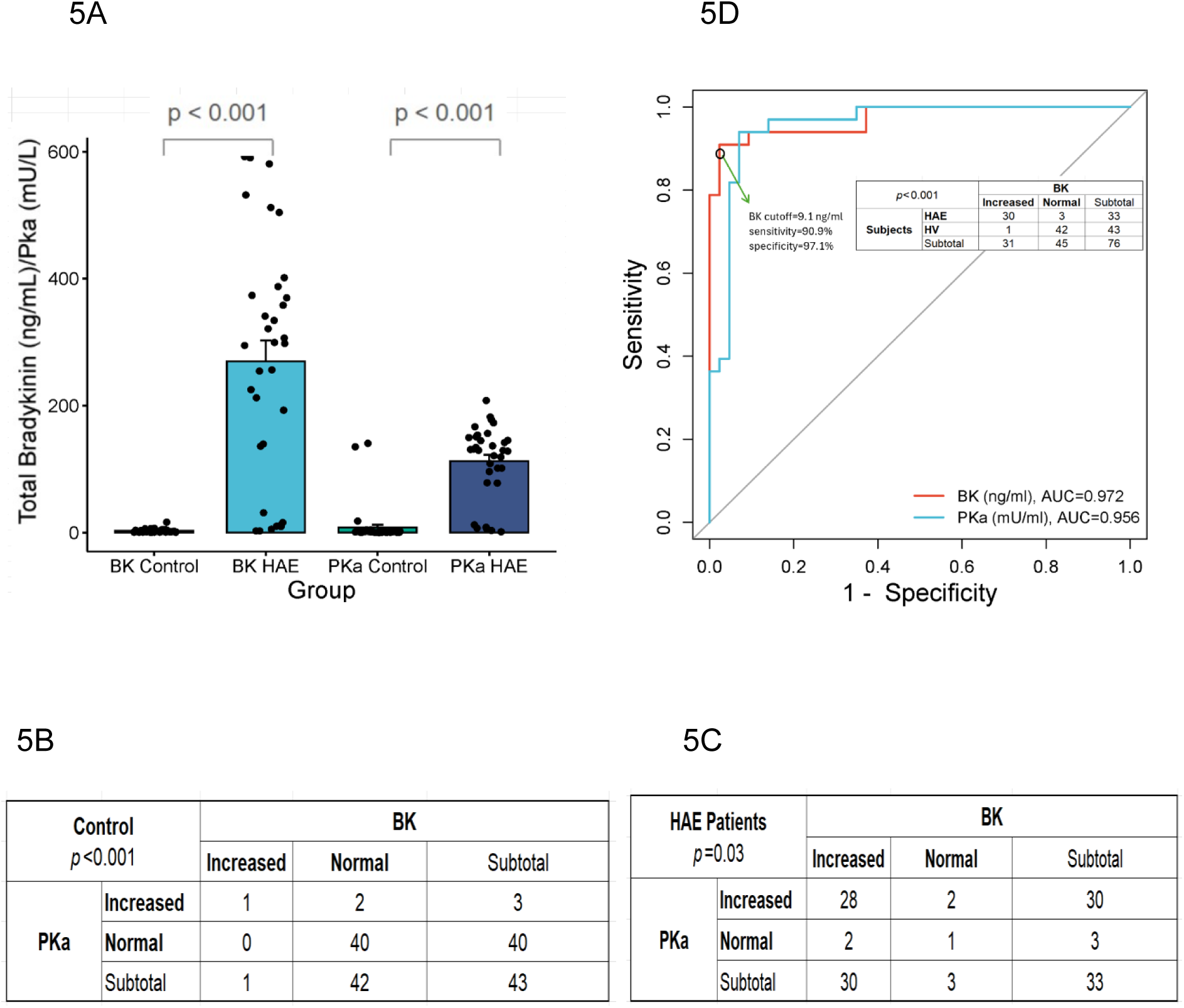
Cold-induced total BK concentrations and plasma kallikrein activities (PKa) in 43 control subjects and 33 HAE-C1INH patients in a validation cohort. (**5A**) Both total BK concentrations and PK activities were significantly increased in HAE subjects. Nearly all samples demonstrated changes in the same direction across control (Figures 5B) and HAE (Figures 5C) groups. Receiver Operating Characteristic (ROC) analysis highlighted strong diagnostic performance for both BK and PKa assays (Figure 5D).

## DISCUSSION

In this study, a quantitative LC-MS/MS method for measurement of BK and its major metabolites was successfully developed and validated in accordance with the regulatory guidelines for both commercial and research laboratories.^17^ We found that cold incubation of whole blood in EDTA without protease inhibitors at 4°C for 1–3 days could preferentially activate the contact system with highly elevated BK levels in HAE-C1INH subjects but not in controls. To the best of our knowledge, this is the first report to apply cold activation to bradykinin measurement for HAE diagnosis and management.

LC-MS/MS offers a tool for the determination of multiple peptides simultaneously, which would allow for the collection of more information and capture a more comprehensive picture of the status of the bradykinin metabolism than the measurement and reporting of single peptides. Our data confirmed the sum of the individual peptides was more representative of the HAE-C1INH pathophysiology (**Figure 3**). This is in agreement with the reports in the literature.^11,13^

Our method differs from previously published LC-MS/MS protocols.^11,12,16^ Instead of protease inhibitors (PI), we used cold activation without PI. The cold activation process produced a greater elevation of total BK (to tens and hundreds ng/mL level) in HAE-C1INH subjects than controls, confirmed by the increased PK activity in the same samples (**Figure 5**). The instability of the contact system in HAE-C1INH is likely the explanation for this phenomenon. The cold activation greatly widened the differences between HAE-C1INH and controls. Although using PI can certainly provide more stability of the KKS (reduction of degradation and generation), the large variability of normal range created a low separation window hindering meaningful clinical application.^11^ The cold activation process increased the BK levels in HAE-C1INH subjects by an average >100 fold (324.3 ± 54.7 ng/mL vs 2.3 ± 0.3 ng/mL; mean±S.E.M., p < 0.001), which essentially eliminated the concern regarding the sample collection related baseline BK level variabilities. It is evidenced by the data that the first and second collection tubes of blood showed no BK differences. The detailed comparisons of PI-based endogenous BK and cold-induced BK detection methods are illustrated in **Table E3** in the Online Repository available at www.jaci-global.org.

HAE-C1INH can be regarded as a prototype of bradykinin mediated angioedema (AE-BK). At this point, no reliable diagnostic tests are available for AE-BK other than C1 and C1INH measurement. Genetic tests can only provide clues for a small proportion of the AE-BK with normal C1INH. Measurement of BK level can potentially provide an additional biomarker for these patients. We will report our clinical data in a companion paper.

In addition to angioedema, BK also plays a role in a wide spectrum of conditions, particularly those associated with inflammation, vascular dysfunction, and pain, such as COVID-19^21^, sepsis.^22^ Use of ACEIs^23^ and estrogen^15^ has been shown to activate the contact system by upregulating components of the KKS, which increases BK production and enhances the sensitivity of BK receptors. Our study excluded those subjects from the controls.

## Conclusion

Bradykinin measurement via LC-MS/MS can be a useful biomarker for HAE-C1INH and other bradykinin involved disease conditions. The instability of contact system in HAE-C1INH can be greatly amplified by cold activation. Using this robust assay, we can measure the differences in BK levels between HAE-C1INH and normal subjects.

## DISCLOSURE STATEMENT

The authors declare no relevant conflicts of interest.

## Abbreviations used

ACEI: angiotensin-converting enzyme inhibitor
AE: angioedema
AUC: area under curve
BK: Bradykinin
C1INH: C1 esterase inhibitor
CAP: College of American Pathologists
CLIA: Clinical Laboratory Improvement Amendments of 1988
HAE-C1INH: hereditary angioedema due to C1INH deficiency/dysfunction
HV: healthy volunteer
HMWK: high-molecular-weight kininogen
IS: internal standard
KKS: kallikrein-kinin system
LDT: Laboratory Developed Test
LC-MS/MS: Liquid Chromatography-Tandem Mass Spectrometry
LLOQ: lower limit of quantification
PI: protease inhibitor
PK: plasma kallikrein
RE: relative error
ROC: receiver operating characteristic
S/N: signal/noise

## Data Availability

All data produced in the present study are available upon reasonable request to the authors.

## Supplemental Material

### MATERIALS AND METHODS

#### LC-MS/MS Method Validation

Following the US FDA bioanalytical validation guidelines, we conducted linearity, accuracy, precision, sensitivity, recovery, matrix effect, carryover, and stability assessment.

Linearity was evaluated using replicates of eight calibrators covering a concentration range from 0.1 ng/mL to 1000 ng/ml. Calibration curve standards were deemed acceptable if the deviation from their nominal concentration (relative error, RE) was within ±15% (±25% at the Lower Limit of Quantification, LLOQ).

Accuracy and precision were established using five replicates from four QC levels across three independent runs on different days. Accuracy was determined as the deviation of the measured concentration from the nominal concentration, both within-run and between-run. The LLOQ was set at 0.1 ng/mL, with a signal-to-noise ratio (S/N) greater than 10:1.

Carryover was assessed by injecting solvent blanks following the highest calibrator injections. Potential interferences were evaluated by spiking common medications and chemicals into blank matrix samples containing known analyte concentrations. The matrix effect was assessed by comparing post-extraction matrices from different subjects, spiked with reference standards, to neat solvent measurements. Stability studies were performed at three concentration levels under varying storage conditions, with sample extraction and measurement conducted at multiple time points.

**Table E1.** HPLC Parameters for BK Separation.

| LC Column | Kinetex Biphenyl Column |
| --- | --- |
| Mobile phase A | 0.1% formic acid in water |
| Mobile phase B | 0.1% formic acid in methanol |
| Flow rate | 0.5 mL/min |
| Run time | 6 min |
| Injection volume | 10 μL |
| Gradient (min) | % Mobile phase B |
| 0.0 | 12 |
| 0.8 | 20 |
| 2.0 | 25 |
| 3.0 | 40 |
| 4.0 | 95 |
| 5.0 | 95 |
| 5.1 | 12 |
| 6.0 (stop time) | 12 |
| Column oven temperature: 40 $^{\circ}$C | |

#### RESULTS

**Figure E1.**
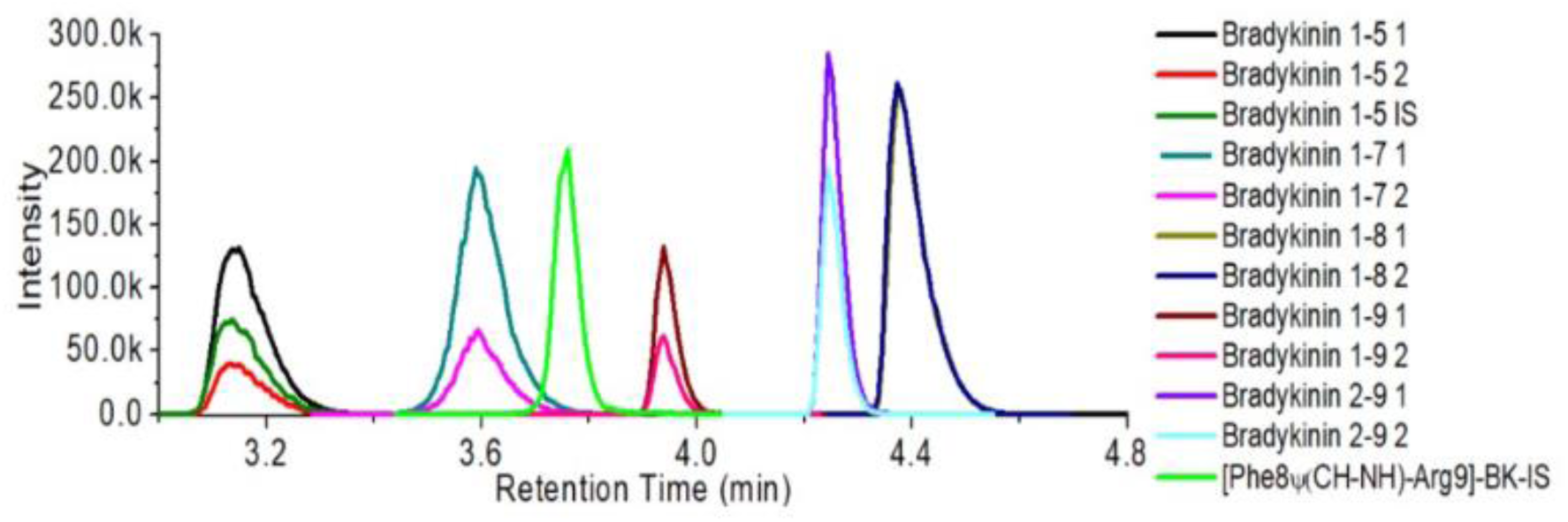
A representative LC-MS/MS chromatogram of Calibrator 3 (0.5 ng/mL for each analyte), illustrating clear separation among all analytes. The signal intensities for all analytes are notably robust, ensuring the reliability and accuracy of our analytical method.

**Table E2.** Assessment of linearity and dynamic range.

| Analyte | Linearity |  |  | Dynamic range |  |  |
| --- | --- | --- | --- | --- | --- | --- |
|  | Mean r | Regression | Weighting | LLOQ (ng/mL) | S/N at LLOQ | ULOQ (ng/mL) |
| BK1-5 | 0.99996 | Linear | 1/x | 0.1 | 51 | 1000 |
| BK1-7 | 0.99960 | Linear | 1/x | 0.1 | 30 | 1000 |
| BK1-8 | 0.99985 | Linear | 1/x | 0.1 | 41 | 1000 |
| BK1-9 | 0.99975 | Linear | 1/x | 0.1 | 19 | 1000 |

LLOQ: lower limit of quantification; ULOQ: upper limit of quantification; S/N: signal-to-noise ratio.

**Table E3.** Different values of PI-based and cold-induced BK detections.

|  | **PI-based endogenous BK**<br>(in the literature) | **Cold-induced BK**<br>(this study) |
| --- | --- | --- |
| **Sample type** | EDTA plasma | EDTA whole blood |
| **Protein inhibitor (PI)** | add PI cocktails during blood collection | no |
| **Activation *in vitro*** | no | cold activation at 4°C for 1-3 days |
| **BK concentration** | Picogram/mL (pg/mL) | Nanogram/mL (ng/mL) |
| **Effect of blood collection process** | can be significant | negligible |
| **Applicability in clinical practice** | limited | more feasible |
| **BK Result Interpretation** | Reflect BK level at the time of blood collection | Reflect the stability of the contact system ex vivo |
| **BK Levels in HAE-C1INH between attacks** | slightly increase or no increase | most subjects showed significant increase |
| **Diagnostic Window** | narrow | wide |

## Notes

### Competing Interest Statement

The authors have declared no competing interest.

### Funding Statement

This study was funded by Foundation for Rare Disease Research and Virant Diagnostics, Inc.

### Author Declarations

The study was conducted following the guidelines of a clinical study protocol approved by a central Virant Institutional Review Board (IRB protocol number: Virant-A0001).

